# How are residents trained in neuropathology? A survey of neurology program directors in the United States

**DOI:** 10.1101/2020.08.16.20176222

**Authors:** Appaji Rayi, Kiran Rajneesh, Vineet Punia, Amanda R. Start

## Abstract

To understand the current state of neuropathology education during neurology residency training in the United States, we electronically distributed a 16-item survey to 150 adult and 70 child neurology program directors (PDs). The survey inquired about their residency program characteristics, neuropathology curriculum and assessment methods, trainee performance in the subject and attitude about neuropathology education. Descriptive analysis was used to summarize categorical variables as frequencies and percentages and continuous as means and standard deviations. We conducted a series of Mann-Whitney U and Fisher’s exact tests to evaluate differences between various program characteristics. Sixty-four (29%) PDs responded to the survey, including 45 (30%) adult and 19 (27%) child neurology PDs. Thirty-one programs required a dedicated neuropathology rotation typically during the latter years of the program. Residency in-service training exam (RITE) was the main assessment tool (92%) for assessing the trainee’s knowledge in neuropathology. Overall, 87% of the PDs agreed that neuropathology is essential and 85% agreed that there is a clear need for a defined neuropathology curriculum during residency training. There was no difference in the RITE scores between programs with and without a dedicated neuropathology rotation. We conclude that a neuropathology rotation was felt to be essential even though the RITE scores did not differ between programs with and without a dedicated rotation. Alternative evaluation methods and neuropathology training techniques such as web modules, virtual reality may be helpful tools to optimize training and need consideration.

## Introduction

Neuropathology is the study of disease of nervous system tissue. A good understanding of neuropathology is critical for both general neurologists and sub-specialists to effectively assess and manage patients with neurological disorders. Neuropathology content typically comprises between 6 and 15% of the annual Residency In-Training Exam (RITE), a standard metric for assessing resident’s educational progress (1). Between 22 and 28% of the questions on the initial licensing exam for neurology are related to neuroscience and mechanism of disease (2). Despite its importance, the Accreditation Council for Graduate Medical Education (ACGME) does not require neuropathology training in the curriculum for neurology residents (3). Presently, individual programs define their own neuropathology curriculum for their trainees. In this study, our goal was to conduct a national survey to assess and understand the current state of neuropathology education during neurology residency training in the United States (US).

## Methods

### Study design and population

We designed a 16-item, online survey to assess a) the characteristics of the neurology residency program, b) the program’s neuropathology curriculum and assessment methods, and c) the program director’s (PD) attitudes about neuropathology education (see Figure 1). We electronically distributed the survey to 150 adult and 70 child neurology PDs using Qualtrics, an online tool for conducting and managing surveys (4). Consistent with the tailored design method(5), we sent each PD up to four email invitations over a 3-week period in May of 2019. Each email provided a brief summary of the study and instructions for accessing the online survey through Qualtrics.

**Figure 1.**
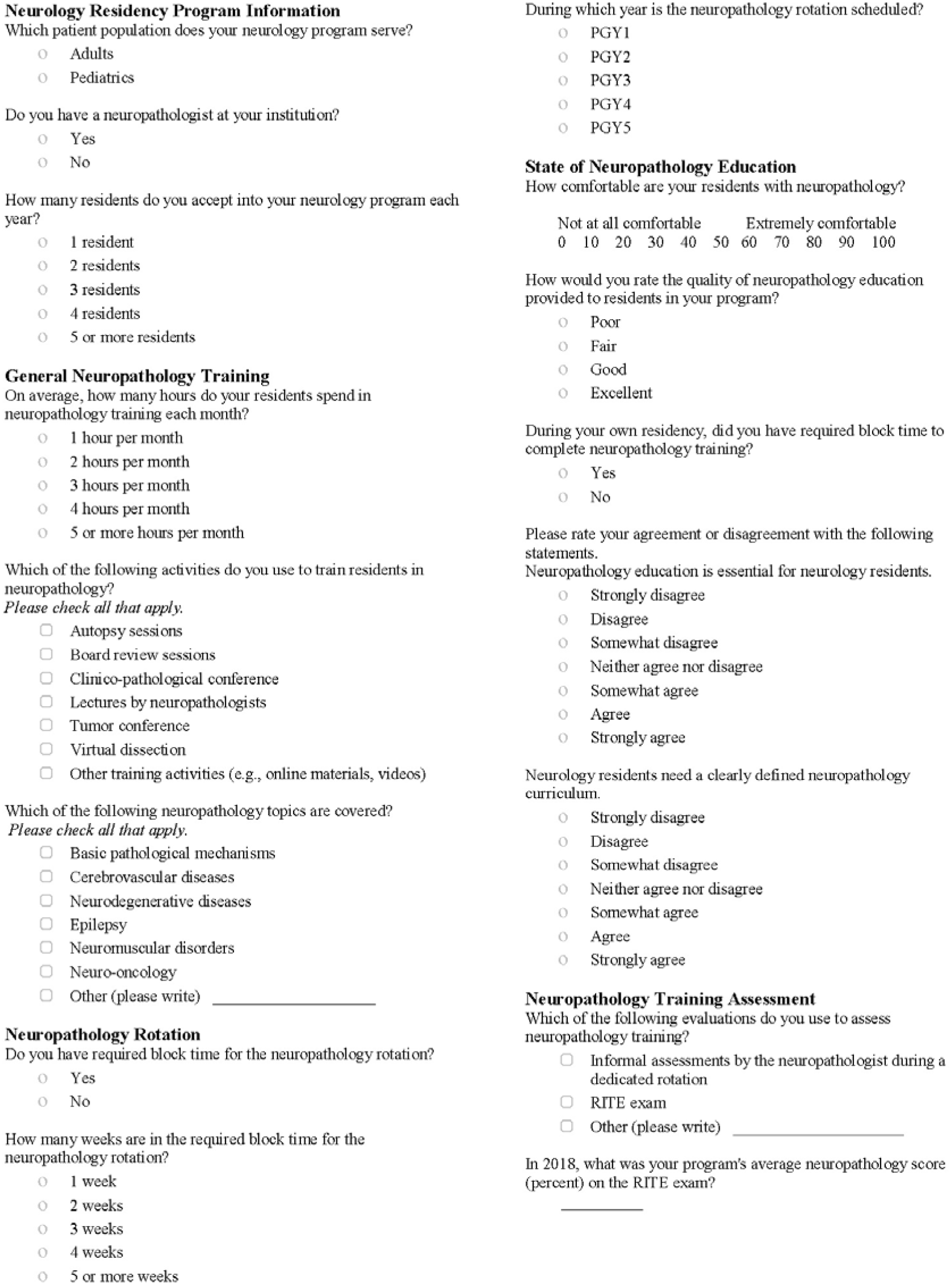
16-Item, Online Survey.

### Standard protocol approvals and consents

The Ohio State University institutional review board (IRB) provided ethical oversight and approval of this study. All participants provided electronic informed consent.

### Statistics

We conducted descriptive analyses for all survey data, summarizing categorical data as frequencies and percentages and continuous data as means and standard deviations. We conducted a series of Mann – Whitney U and Fisher’s exact tests to evaluate differences between a) adult and child programs and b) programs with and without a required neuropathology block rotation. We analyzed the data using SPSS version 25 (6).

### Data availability

Anonymized data is available upon request from the Corresponding Author.

## Results

Sixty-four (29%) PDs responded to the survey, including 45 (30%) adult PDs and 19 (27%) child neurology PDs.

### Program characteristics

The majority of programs (n = 60; 90%) had a neuropathologist on staff. More than half (n = 39; 61%) of programs admitted 5 or more residents per year (see table 1). Child programs tended to have fewer residents than adult programs (U = 175.00, p < 0.001; see Table 1).

**Table 1.** Characteristics of the training program, neuropathology rotation, neuropathology curriculum and trainee assessment

| **Item** | ***n*** | **Frequency (%)** | **Mean (SD)** |
| --- | --- | --- | --- |
| **Patient population served** | 64 |  |  |
| **Adults** |  | 45 (70.31) |  |
| **Pediatrics** |  | 19 (29.69) |  |
| **No. residents accepted per year** | 64 |  |  |
| **1 resident** |  | 2 (3.13) |  |
| **2 residents** |  | 10 (15.63) |  |
| **3 residents** |  | 7 (10.94) |  |
| **4 residents** |  | 6 (9.38) |  |
| **5 or more residents** |  | 39 (60.94) |  |
| **Residency year for required neuropathology block** | 31 |  |  |
| **PGY1** |  | 0 (0.00) |  |
| **PGY2** |  | 1 (3.23) |  |
| **PGY3** |  | 8 (25.81) |  |
| **PGY4** |  | 18 (58.06) |  |
| **PGY5** |  | 4 (12.90) |  |
| **Hours of training** | 58 |  |  |
| **Up to 1 hour per month** |  | 37 (63.79) |  |
| **2 hours per month** |  | 9 (15.52) |  |
| **3 hours per month** |  | 5 (8.62) |  |
| **4 hours per month** |  | 7 (12.07) |  |
| **Subtopics in the neuropathology education** | 63 |  | 4.71 (1.65) |
| **Basic pathological mechanisms** |  | 41 (65.08) |  |
| **Cerebrovascular** |  | 49 (77.78) |  |
| **Neurodegenerative diseases** |  | 56 (88.89) |  |
| **Epilepsy** |  | 38 (60.32) |  |
| **Neuromuscular disorders** |  | 50 (79.37) |  |
| **Neuro-oncology** |  | 54 (85.71) |  |
| **Other<sup>a</sup>** |  | 9 (14.29) |  |

<sup>a</sup> Other topics identified included, analysis of specimens, developmental and migrational disorders, embryonic development, gross anatomy, and neuro-infectious diseases.

### Neuropathology rotation

Although 75% (n = 46) of PDs had required block time for neuropathology during their own residency training, only half (n = 32; 53%) of the programs currently require a block time for the neuropathology rotation. A greater proportion of child neurology programs required block time (n = 14; 82%) compared to adult programs (n = 18; *p* = 0.004).

When required, the rotation was most frequently 4 weeks in length (n = 19; 59%), followed by 2 weeks (n = 6; 19%) and 5 or more weeks (n = 5; 16%; see table1). The rotation was typically required during the fourth (n = 18; 58%) or third (n = 8; 26%) year of training.

### Neuropathology curriculum

Most of the programs (n = 37; 64%) spent 1 hour or less on neuropathology education per month (see Table 1). The average program provided training on 4.71 (SD = 1.65) of 7 subtopics in neuropathology (e.g., basic pathological mechanisms; cerebrovascular; neurodegenerative diseases). Neuropathology education was most commonly disseminated through board review sessions (n = 45; 71%), neuropathologist lectures (n = 44; 70%), and clinico-pathological conferences (CPC; n = 35; 56%).

### Trainee assessment in neuropathology

On average, PDs reported their trainees were somewhat uncomfortable with neuropathology (on a scale from 0 [not at all comfortable] to 100 [extremely comfortable]; M = 41.18; SD = 22.47). However, PDs of programs *with* a required neuropathology block (M = 50.07, SD = 21.61) reported greater resident comfort than PDs of programs *without* a required neuropathology block (M = 30.92; SD = 19.11; U = 203.00, p = 0.002).

The primary method of trainee assessment in neuropathology was the RITE exam (n = 59; 92%), although informal assessments by a neuropathologist (n = 26; 41%) were also common. The mean neuropathology score on the RITE exam in 2018 was 57.48 (SD = 10.87); however, only 62% (n = 40) of respondents provided RITE exam data. There was no difference in RITE scores between programs *with* (M = 58.05; SD = 7.98) and *without* a dedicated neuropathology block (M = 56.84, SD = 13.57; U = 168.50, p = 0.41).

### PD perceptions and attitudes

Most of the PDs felt the quality of their program’s neuropathology education was fair to good (n = 40, 66%; M = 2.56, SD = 0.98). Perceived program quality varied by the requirement of a neuropathology block. PDs of programs *with* a required neuropathology block (M = 3.00, SD = 0.88) perceived their programs to be of higher quality than PDs of programs *without* a required neuropathology block (M = 2.07, SD = 0.84; U = 216.50, p < 0.001).

Overall, the majority of PDs (n = 53; 87%) agreed that neuropathology education is essential for neurology residents (M = 5.70, SD = 1.30). The majority (n = 52; 82%) also agreed that there is a clear need for a defined neuropathology curriculum for residents (M = 5.67, SD = 1.45).

## Discussion

The majority of US adult and child neurology PDs agree that neuropathology training is essential for neurology residents and that there is a clear need for a defined neuropathology curriculum. This suggests that the importance of understanding neuropathology during training is well recognized by the neurology education leadership and in fact, a structured, formal education in neuropathology is highly desired by them. The neurosurgical training programs in the United States have a dedicated neuropathology rotation in their curriculum as set forth by the ACGME (7). This underscores the importance of training in the subject realized by our colleagues in neurosurgery. Over 75% of the PDs had a dedicated neuropathology rotation during their own training compared to 52% in their present programs. Although, the study was not actually designed to compare the state of current neuropathology training with the by gone years, this finding certainly shows a trend towards decreasing formal and structured neuropathology training over years. Multiple factors affect the implementation of a dedicated rotation in to the residency program. These may include limited time, increased number of rotations due to specialization of neurology subspecialties, lack of availability of standardized modules and teaching material, access to allied rotations of neurosurgery and/or neuroradiology which may be deemed as an adequate exposure to neuropathology. However, further research is needed to assess these proposed factors.

Another interesting observation in this study is that the size of the training program did not correlate with dedicated rotation to neuropathology. Child programs have more dedicated rotation to neuropathology, which is contrary to popular belief of limitations of personnel and time. The exact factors that determine the need or resources for dedicated neuropathology rotation at the individual program level needs further study.

Monitoring RITE score remains the main method of evaluating resident’s performance in the subject. Although the scores did not differ between program based on the presence or absence of a dedicated neuropathology rotation, the perception of quality of the neuropathology education and comfort level of the residents in the subject by the PDs were significantly different. These differences could be due to other ways of assessment including evaluation by neuropathologists and resident feedback. Conversely, it could also imply that formal neuropathology training does not lead to any objective difference in performance on the RITE exam and the significant difference perceived by PDs is biased by the dedicated resources devoted in their program for the neuropathology education.

The neuropathology curriculum was primarily disseminated through neuropathology board review sessions, lectures by a neuropathologist, tumor boards and CPCs. Other innovative methods such as virtual reality-based education and online sources are also used. The duration of the rotation is variable and majority of the programs have a 4-week long block rotation, which is probably appropriate and are mostly completed during the final year of training. We speculate that completing the neuropathology rotation during the initial years of training may be appropriate as well, but doing the core clinical rotations earlier in the training hold higher prominence and are usually mandatory.

There are few limitations that need consideration. The response rate appears to be low which might limit the generalizability of the findings. However, a 30 to 40 % response rate is typical for surveys of similar cohorts. (8, 9) And the proportion of adult and child neurology programs in our sample (70 % and 30 % respectively) is representative of the proportion of adult and child neurology programs in the United States (68% and 32 % respectively). Another limitation is the self-report nature of the data. However, following a tailored design method, several survey design strategies were implemented to increase the likelihood of accurate reporting. (5)

## 5. Conclusion

This study elucidates the current state of neuropathology training in neurology residency training in the United States. We hope that our study will help initiate a conversation about formalizing training in neuropathology. Conducting a similar survey of the current trainees in neurology might be useful to evaluate the assessment methods of their knowledge in neuropathology, methods of learning/teaching employed, interest in the subject and any educational barriers. Considering alternative neuropathology training techniques such as web modules, virtual reality may be helpful tools to optimize training exposure to essential subspecialty topics of fundamental value like neuropathology within the current time constraints.

## Data Availability

Anonymized data is available upon request from the Corresponding Author.

## Acknowledgements

We thank Mr. Noah Wren (The Ohio State University) for his diligent work in preparing the manuscript tables and figures. We also thank all of the adult and child neurology program directors who invested their time to complete the survey, thereby contributing to our understanding of neuropathology training.

## Disclosures

Dr. Rayi reports no disclosures.

Dr. Rajneesh reports no disclosures.

Dr. Punia reports no disclosures.

Dr. Start reports no disclosures.

## Declaration of Competing Interest

The authors declare that they have no known competing financial interests or personal relationships that could have appeared to influence the work reported in this paper.

## Notes

### Competing Interest Statement

The authors have declared no competing interest.

### Funding Statement

None

### Author Declarations

The Ohio State University IRB has provided ethical oversight and approval of this study.

